# Development and external validation of deep learning models for spontaneous preterm birth prediction from mid-trimester cervical ultrasound

**DOI:** 10.64898/2026.07.17.26358221

**Authors:** Radhika Chanian, Divyanshu Mishra, Rahul Jain, Nikhil Sharma, Ashok Khurana, Reva Tripathi, Abhinav Jain, GARBH-Ini study group, Nitya Wadhwa, J. Alison Noble, Ramachandran Thiruvengadam, Bapu Koundinya Desiraju, Shinjini Bhatnagar

**Affiliations:** Maternal and Child Health Program, Translational Health Science and Technology Institute, Faridabad, India; The Ultrasound Lab, New Delhi, India; Department of Obstetrics & Gynecology, Hamdard Institute of Medical Sciences and Research and Hakeem Abdul Hameed Centenary Hospital, New Delhi, India; Department of Radiodiagnosis, Hamdard Institute of Medical Sciences and Research and Hakeem Abdul Hameed Centenary Hospital, New Delhi, India; Institute of Biomedical Engineering, University of Oxford, United Kingdom

**Keywords:** preterm birth, cervical ultrasound, prediction model, external validation, deep learning

## Abstract

Preterm birth is the leading cause of neonatal death. Despite sustained efforts to identify high-risk women in the mid-trimester, accurate prediction remains difficult. Quantitative cervical ultrasound texture has been proposed as a predictor of spontaneous preterm birth. However, earlier models were developed in small single-centre samples and were not externally validated. We developed image-texture (Local Binary Patterns with a Random Forest), deep-learning (Vision Transformer), clinical-variable, and multimodal models to predict spontaneous preterm birth on the prospective GARBH-Ini cohort. We then externally validated our best models on an independent cohort scanned on a different ultrasound machine. Our best overall model reached an internal-test area under the receiver-operating-characteristic curve of 0.71 (95% CI 0.60, 0.82), but performed modestly at 0.52 (95% CI 0.38, 0.64) externally. The deep-learning and multimodal models did not perform better. Discrimination appeared higher in a clinically high-risk subgroup at the 34-week threshold. These estimates were imprecise because of few cases and need to be confirmed in future studies. Among the several likely reasons for the modest external performance is the heterogeneity of preterm birth. Predicting distinct preterm-birth subtypes separately, and integrating additional biomarkers and data domains, might improve model performance.

## Introduction

Preterm birth, defined as birth before 37 completed weeks of gestation, remains the leading cause of neonatal death and of mortality in children younger than 5 years, and an estimated 13.4 million infants were born preterm worldwide in 2020, a figure that has not measurably declined over the previous decade^1,2^. Preterm birth is not a single disease but a syndrome, arising through diverse maternal, fetal, placental, and parturition pathways, and this etiological heterogeneity is widely recognized in the phenotypic classification of the condition^3^. Identifying women at risk early enough to intervene is therefore of clinical importance but is complicated by the multiple distinct mechanisms that lead to a common outcome.

The burden of preterm birth is concentrated in southern Asia and sub-Saharan Africa^1,4^, and in India approximately 13% of births are preterm. Current mid-trimester predictors have limited accuracy in asymptomatic, low-risk women: cervical length has modest sensitivity and is subject to inter-observer and intra-observer variation, and biochemical markers such as fetal fibronectin add little in this population^5–7^. An accurate screening tool that could be applied to the routine mid-trimester scan would address a clear gap in antenatal care.

Quantitative analysis of cervical ultrasound texture, and more recently deep-learning models applied to cervical images, have been proposed to extract preterm-birth-related signal beyond cervical length, with reported areas under the curve of 0.77 to 0.84^8–13^. These studies were, however, conducted in small, single-centre samples and were not tested on an independent cohort or a different ultrasound machine. Deep-learning models applied to medical images are known to degrade when the acquisition device or setting changes, a phenomenon termed scanner domain shift^14^, yet no cervical-image preterm-birth model has been subjected to external validation under these conditions.

We set out to develop a universal mid-trimester screening model for spontaneous preterm birth from cervical ultrasound. We developed image-texture, deep-learning, clinical, and multimodal models on a large prospective cohort and externally validated our best models on an independent prospective cohort scanned on a different machine. We evaluated whether cervical texture adds predictive signal over cervical length, clinical variables, and deep-learning models. We assessed performance separately in the overall and clinically high-risk subgroups, and at the 37-week and 34-week thresholds.

## Results

We assessed eligibility and derived the analytic samples for both cohorts (Figure 1). Exclusions were made for multiple births (N=44), stillbirths (N=177), abortions (N=210), and loss to follow-up with missing outcomes (N=773) or missing scans (N=1582). In GARBH-Ini, we additionally excluded images that could not be retrieved through the de-identification pipeline (N=555). In CORE cohort, exclusions were made for multiple births (N=0), stillbirths (N=3), abortions (N=23), and loss to follow-up with missing outcomes (N=4) or missing scans (N=210).

**Figure 1:**
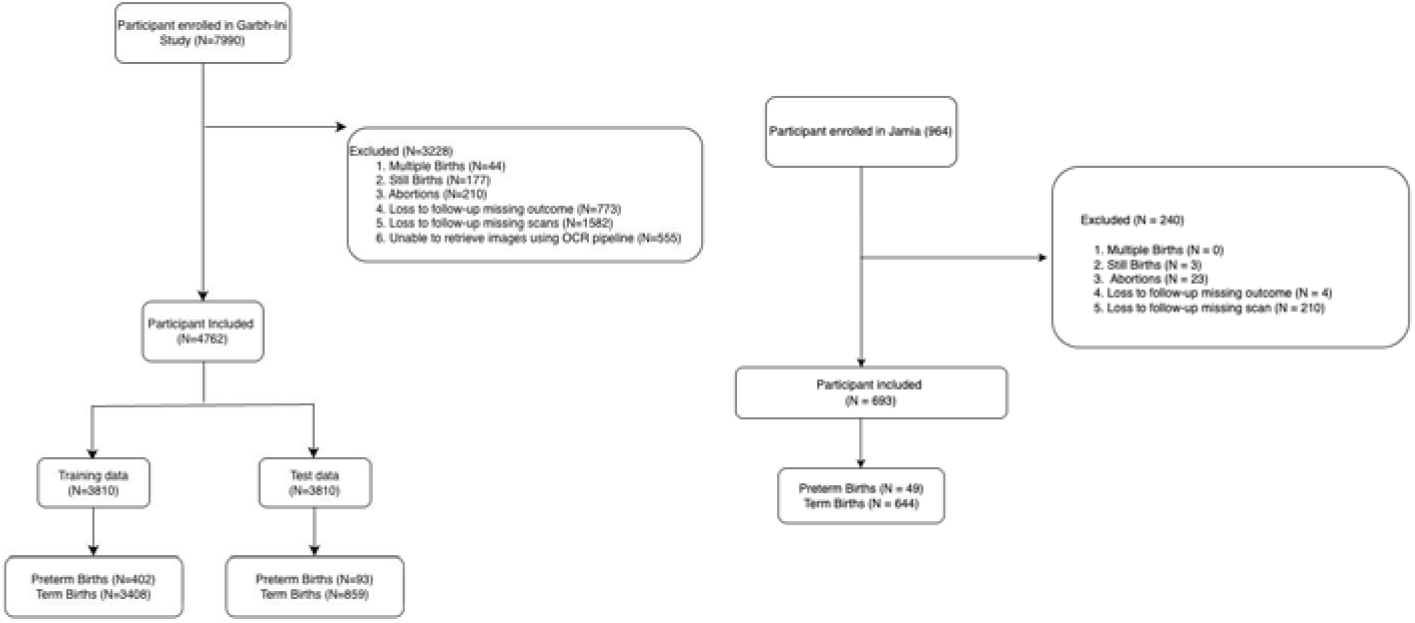
Flow of participants through the two cohorts: (A) the GARBH-Ini development and internal-test cohort (Gurugram Civil Hospital) and (B) the CORE external-validation cohort (Hamdard Institute of Medical Sciences and Research, New Delhi). For each cohort the diagram shows the number assessed, the numbers excluded by reason, and the analytic sample of mid-trimester (18 to 20 week) cervical ultrasound images with a recorded delivery outcome, split into term and spontaneous preterm birth before 37 weeks.

The two cohorts differed in maternal and socioeconomic characteristics (Table 1). GARBH-Ini participants were younger (median age 23.0 vs 27.6 years) and more often underweight (26% vs 7.1%) and were more socioeconomically disadvantaged. The CORE cohort carried a higher overweight (11% vs 32%) and obese (1.8% vs 11%) burden. Several clinical variables were not collected in the CORE cohort.

**Table 1:**
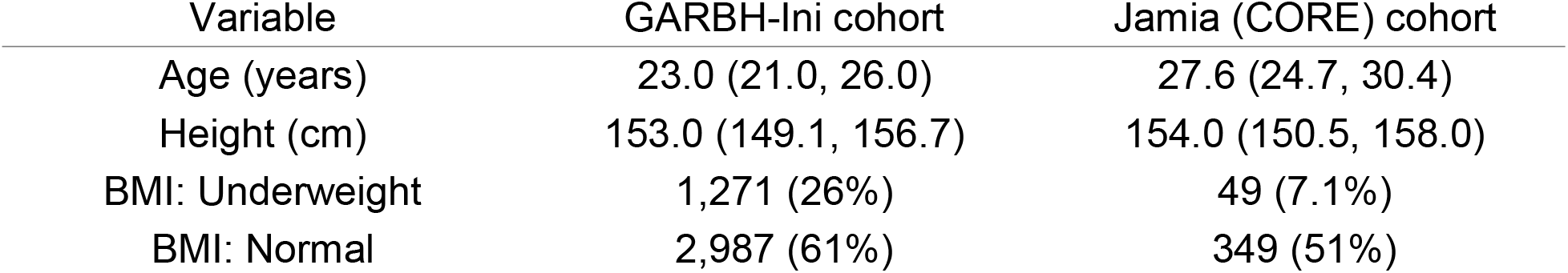
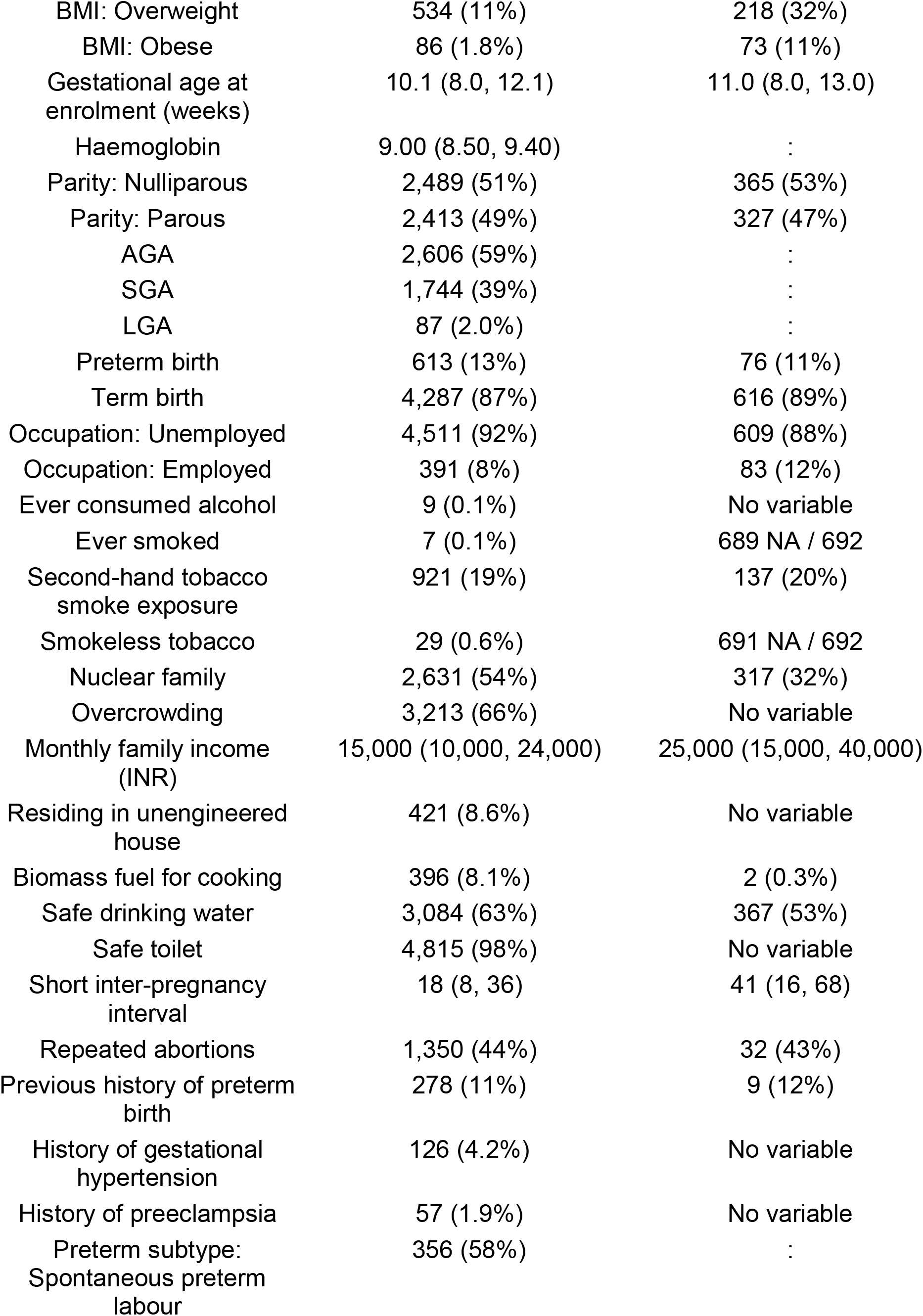
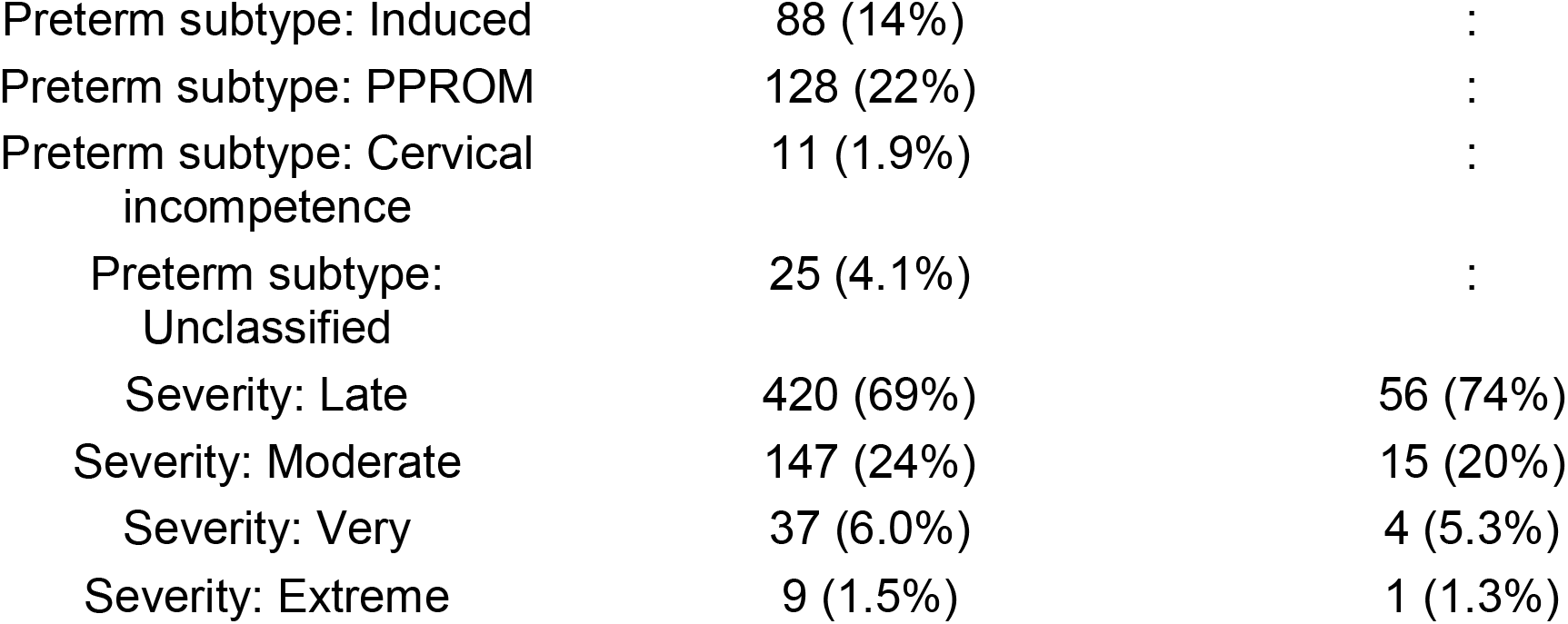
Baseline characteristics of the GARBH-Ini development cohort and the Jamia (CORE) external-validation cohort. Continuous variables are summarised as median (IQR) and categorical variables as n (%). “No variable” or “NA” indicates a field not collected in the Jamia cohort. Some denominators in the source draft exceed the stated analytic N and require reconciliation before submission.

The best-performing image model on internal testing used cervical texture. The cervical region of interest was first manually segmented (Figure 2). Local Binary Patterns were then extracted within this region, and the resulting texture histogram was used as the input to a Random Forest classifier. In the overall group, the cervical-texture Random Forest reached an internal-test AUROC of 0.71 [95% CI 0.60–0.82] (94 images: 12 preterm and 82 term). It fell to 0.52 [95% CI 0.38–0.64] on the external cohort (69 images: 33 preterm and 36 term), indicating discrimination close to chance after a change of cohort and machine (Table 2, Figure 3). The Vision Transformer reached an internal-test AUROC of 0.54 [95% CI 0.46–0.60] (952 images: 93 preterm and 859 term) and an external AUROC of 0.59 [95% CI 0.49–0.68] (578 images: 42 preterm and 536 term). The Vision Transformer and the multimodal models did not outperform the texture model on either test set, and adding cervical length or clinical variables to the texture features did not improve overall discrimination.

**Table 2:** Operating-point performance in the overall group on the internal (GARBH-Ini) and external (CORE/Jamia) test sets, at the selected classification threshold: positive predictive value (PPV), negative predictive value (NPV), specificity, sensitivity, and AUROC, each as point estimate with 95% CI; preterm and term event counts are given with each model.

| Test set | Model (events) | PPV | NPV | Specificity | Sensitivity | AUROC |
| --- | --- | --- | --- | --- | --- | --- |
| Internal | Cervix US images (preterm 93, term 859) | 0.02<br>(0.01, 0.02) | 0.98<br>(0.98, 0.99) | 0.34<br>(0.33, 0.35) | 0.65<br>(0.53, 0.76) | 0.54<br>(0.46, 0.60) |
| Internal | LBP with ROI (preterm 12, term 82) | 0.20<br>(0.08, 0.36) | 0.92<br>(0.84, 0.98) | 0.66<br>(0.57, 0.76) | 0.58<br>(0.26, 0.89) | 0.71<br>(0.60, 0.82) |
| External | Cervix US images (preterm 42, term 536) | 0.01<br>(0.01, 0.02) | 0.99<br>(0.99, 1.00) | 0.59<br>(0.57, 0.61) | 0.50<br>(0.27, 0.72) | 0.59<br>(0.49, 0.68) |
| External | LBP with ROI (preterm 33, term 36) | 0.51<br>(0.37, 0.64) | 0.83<br>(0.50, 1.00) | 0.13<br>(0.03, 0.27) | 0.97<br>(0.90, 1.00) | 0.52<br>(0.38, 0.64) |

**Figure 2:**
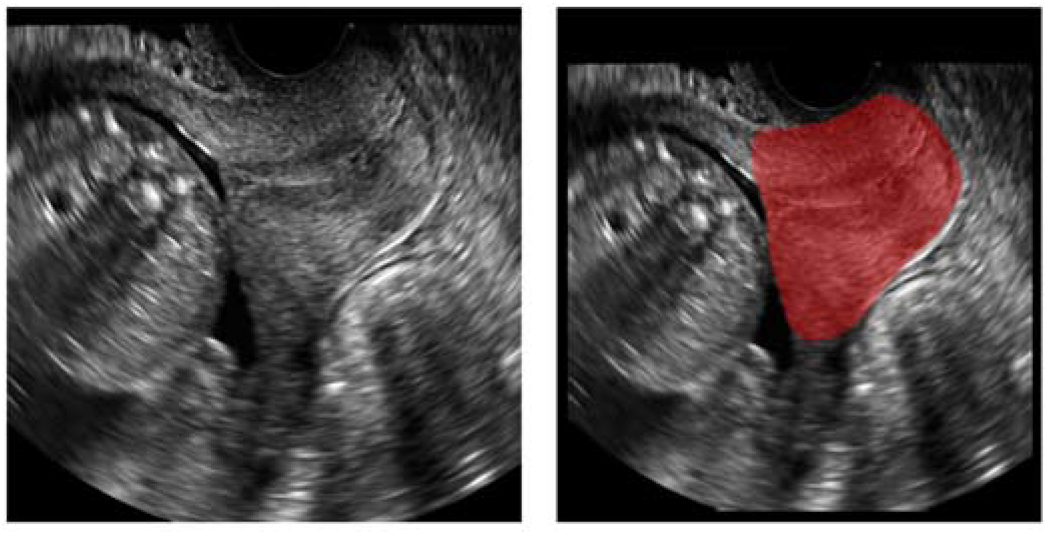
Cervical region of interest used for quantitative texture analysis. (a) A clean mid-trimester transvaginal cervical ultrasound image. (b) The same image with the entire cervical region manually segmented as the region of interest. Local Binary Pattern texture is computed within this mask and summarised as a per-image histogram, the input feature to the Random Forest model.

**Figure 3:**
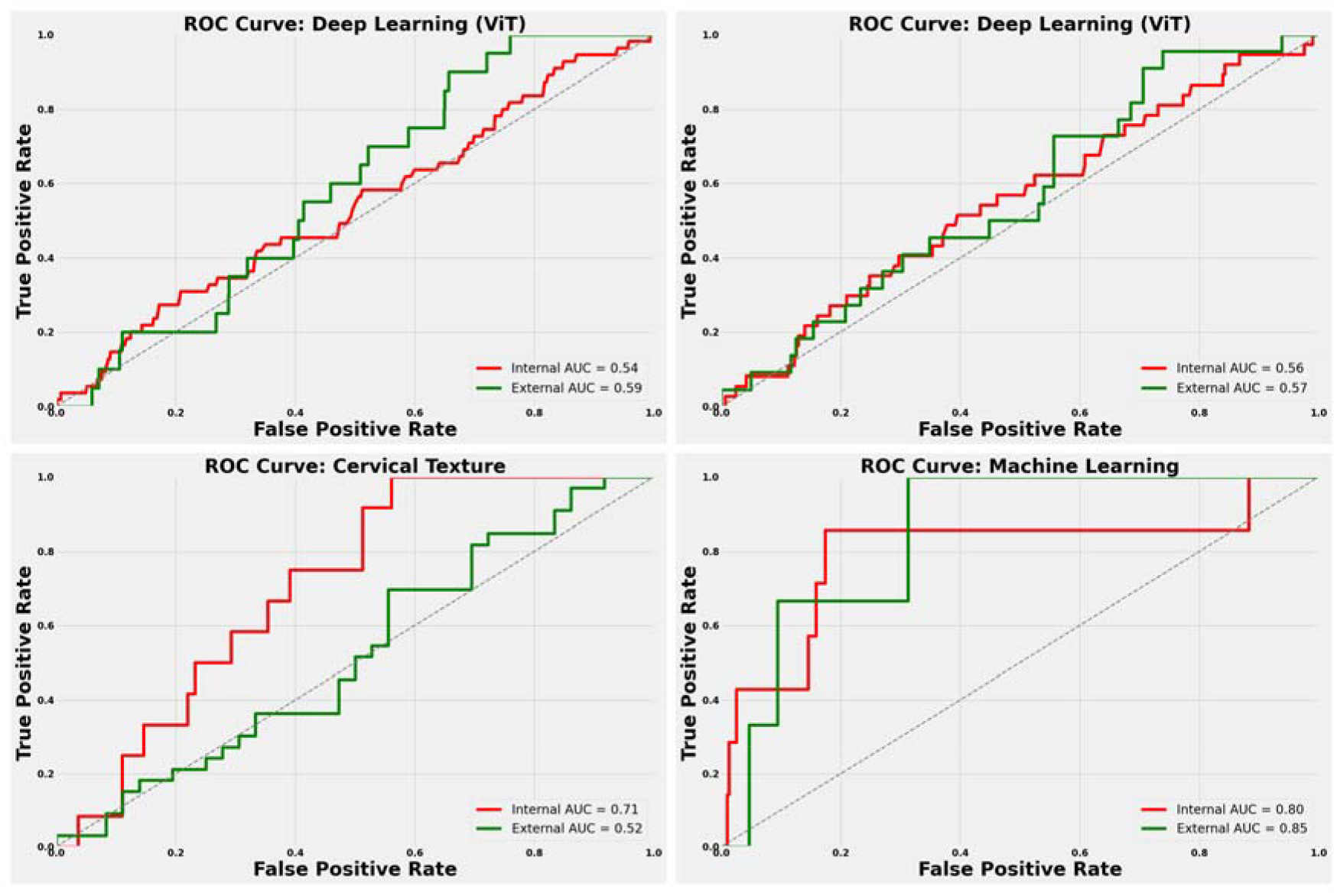
Receiver-operating-characteristic curves on the internal (GARBH-Ini) and external (CORE) test sets. (a) Vision Transformer on cervical images in the overall group (internal AUROC 0.54, external 0.59). (b) Vision Transformer on cervical images in the high-risk group (internal AUROC 0.56, external 0.57). (c) Cervical-texture Random Forest in the overall group (internal 0.71, external 0.52). (d) Machine-learning clinical-plus-cervical-length model in the high-risk subgroup (internal 0.80, external 0.85). The diagonal indicates chance (AUROC 0.50). Internal discrimination for the cervical-texture model falls toward chance externally, whereas the high-risk clinical-plus-cervical-length model remains high on small numbers of events.

Across the full grid of models in the overall group, AUROC values were modest internally and did not generalize externally (Table 2). Due to the low prevalence of preterm birth in these cohorts, all models showed low positive predictive values across the evaluated thresholds (Table2, Table 3).

**Table 3:**
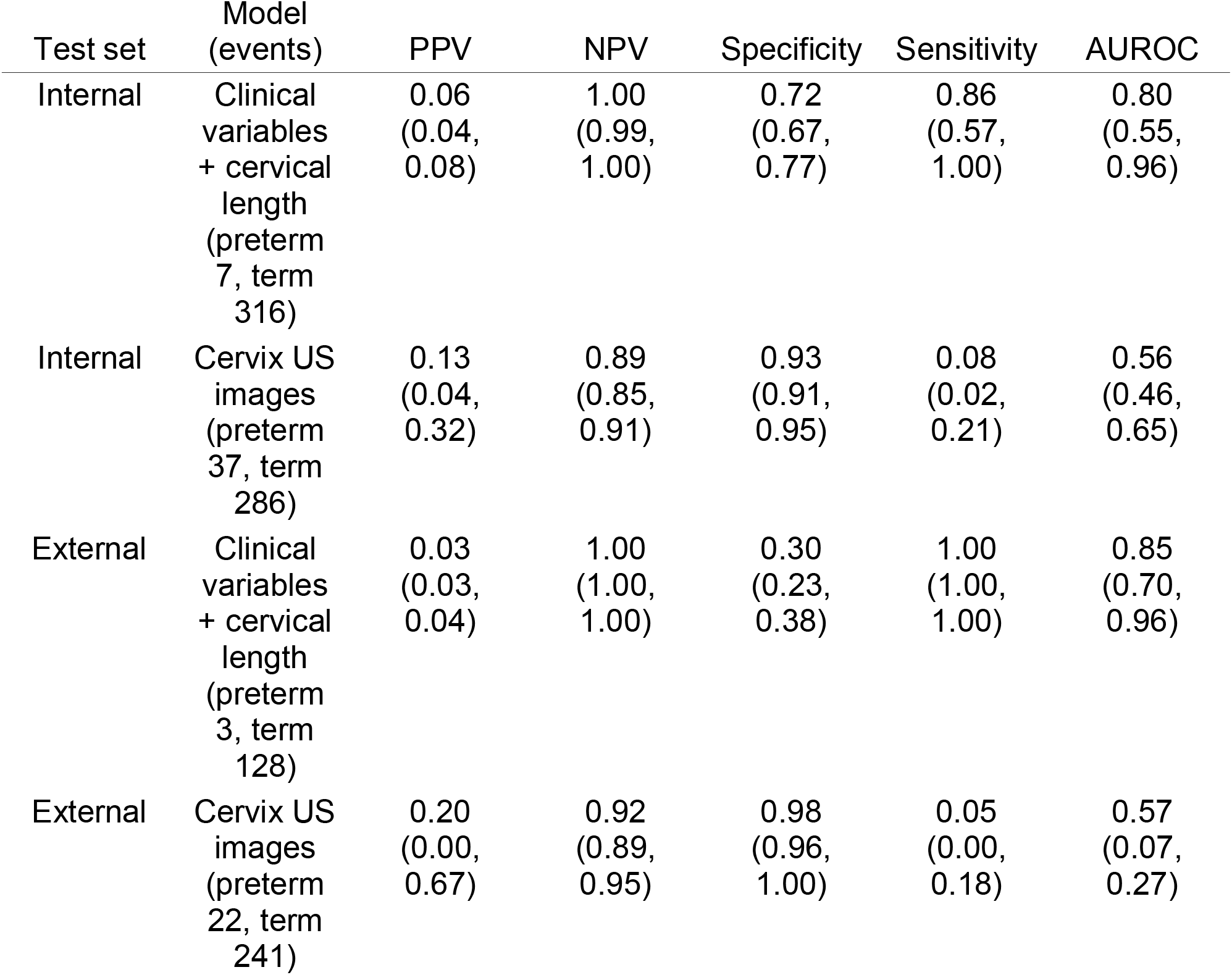
Operating-point performance in the high-risk subgroup on the internal (GARBH-Ini) and external (CORE/Jamia) test sets, at the selected classification threshold: PPV, NPV, specificity, sensitivity, and AUROC, each as point estimate with 95% CI; preterm and term event counts are given with each model. These rows rest on small numbers of preterm events (external as few as three), producing wide CIs and, in places, degenerate operating points (e.g., sensitivity 1.00 at low specificity); they are reported for transparency and should not be read as stable performance estimates. The external cervix-US AUROC CI (0.07, 0.27) is internally inconsistent with its point estimate in the source and requires correction.

| Test set | Model<br>(events) | PPV | NPV | Specificity | Sensitivity | AUROC |
| --- | --- | --- | --- | --- | --- | --- |
| Internal | Clinical<br>variables<br>+ cervical<br>length<br>(preterm<br>7, term<br>316) | 0.06<br>(0.04,<br>0.08) | 1.00<br>(0.99,<br>1.00) | 0.72<br>(0.67,<br>0.77) | 0.86<br>(0.57,<br>1.00) | 0.80<br>(0.55,<br>0.96) |
| Internal | Cervix US<br>images<br>(preterm<br>37, term<br>286) | 0.13<br>(0.04,<br>0.32) | 0.89<br>(0.85,<br>0.91) | 0.93<br>(0.91,<br>0.95) | 0.08<br>(0.02,<br>0.21) | 0.56<br>(0.46,<br>0.65) |
| External | Clinical<br>variables<br>+ cervical<br>length<br>(preterm<br>3, term<br>128) | 0.03<br>(0.03,<br>0.04) | 1.00<br>(1.00,<br>1.00) | 0.30<br>(0.23,<br>0.38) | 1.00<br>(1.00,<br>1.00) | 0.85<br>(0.70,<br>0.96) |
| External | Cervix US<br>images<br>(preterm<br>22, term<br>241) | 0.20<br>(0.00,<br>0.67) | 0.92<br>(0.89,<br>0.95) | 0.98<br>(0.96,<br>1.00) | 0.05<br>(0.00,<br>0.18) | 0.57<br>(0.07,<br>0.27) |

Discrimination appeared higher in the clinically high-risk subgroup, particularly at the 34-week threshold. There, the clinical-variable model reached an internal AUROC of 0.77 [95% CI 0.53–0.95] and an external AUROC of 0.86 [95% CI 0.75–0.98]. The clinical-plus-cervical-length model reached an internal AUROC of 0.80 [95% CI 0.55–0.95] and an external AUROC of 0.85 [95% CI 0.69–0.96] (Table 2, Figure 3). These estimates rest on very small numbers of preterm events: in the high-risk subgroup the clinical-plus-cervical-length analysis included 7 preterm of 323 internally and only 3 preterm of 131 externally. The confidence intervals are correspondingly wide, and several operating points are degenerate (for example, sensitivity 1.00 at low specificity).

The gain in the high-risk subgroup came from the clinical models rather than from the image-based model. The clinical-variable and clinical-plus-cervical-length models reached AUROCs of 0.80 to 0.86 at the 34-week threshold. In the same subgroup, the cervix ultrasound image model discriminated only weakly (Table 3). On the internal test set (323 participants: 37 preterm and 286 term), the image model reached an AUROC of 0.56 [95% CI 0.46–0.65]. On the external test set (263 participants: 22 preterm and 241 term), it reached an AUROC of 0.57 [95% CI 0.07–0.27] (Table 3). The high-risk signal was therefore driven by clinical variables and cervical length, not by cervical imaging. Operating-point performance for the reported models, in the overall group (Table 2) and the high-risk subgroup (Table 3), is shown for the internal and external test sets, with the contributing preterm and term counts given for each row.

## Discussion

We set out to build a universal cervical-ultrasound screening model for spontaneous preterm birth and found that this was not achievable: a cervical-texture model that discriminated moderately on internal testing (AUROC 0.71) fell to chance on an independent cohort scanned on a different machine (0.52), and deep-learning and multimodal models did not perform better. A higher discrimination was observed within a clinically defined high-risk subgroup (AUROC up to 0.86 at the 34-week threshold), but this signal rests on very few events and is exploratory.

Several factors plausibly contribute to the failure of the overall model to generalize externally. The two cohorts differ in case-mix and socioeconomic profile (Table 1) and probably in outcome prevalence, so spectrum shift between development and validation populations is one likely contributor. In addition, cervical ultrasound images vary substantially within and between machines, and image-based deep-learning models are known to degrade under such scanner domain shift^14^; the change of ultrasound vendor between cohorts is therefore a second likely contributor. We cannot separate these explanations with the present data. Either way, the result tempers the earlier, smaller studies that reported areas under the curve of 0.77 to 0.84 for cervical texture without external validation^8–10^: in a larger, prospectively recruited, externally validated setting we did not reproduce that level of performance for a universal model.

That the predictive signal from the cervical-texture model appeared concentrated in the high-risk subgroup is consistent with preterm birth being a heterogeneous syndrome rather than a single condition^3^. We interpret this as support for defining clinically or biologically distinct subtypes and predicting each separately, rather than fitting one universal classifier across all phenotypes; data-driven analyses have similarly resolved preterm birth into distinct subtypes with different correlates^15,16^. We emphasize, however, that our high-risk results are based on as few as three external preterm events and require confirmation in a larger high-risk sample before any clinical inference is drawn.

The second implication is that a single imaging domain is unlikely to be sufficient for a syndrome with diverse biological pathways. Predictive signal for preterm birth has been demonstrated in the cervicovaginal microbiome and metabolome, in maternal proteomic and lipidomic profiles, in cell-free RNA, and in integrated multi-omic models, including in low- and middle-income settings^17–21^. Integrating imaging with clinical, microbiome or metabolome, and genomic data (building on multi-omic work in this population^22^) is, in our view, the more promising direction than refining a single image-based screen.

The major strength of this study is that it is, to our knowledge, among the first cervical-image preterm-birth models to be externally validated on an independent prospective cohort scanned on a different ultrasound machine, reported in full following TRIPOD+AI. Several limitations need to be noted. The external high-risk analyses rest on very small numbers of preterm events, producing wide confidence intervals and unstable operating points, so the high-risk discrimination should be read as exploratory. Because we applied no multiplicity correction across the grid of models, subgroups, and thresholds, we regard the high-risk results as hypothesis-generating rather than as established performance. With few preterm events relative to the number of candidate predictors, the internal estimates are likely optimistic. We did not present calibration estimates or a formal clinical-utility (net-benefit) analysis in this report; given the low positive predictive values observed, the models are not suitable for standalone clinical use in their current form. We did not undertake a formal fairness audit, despite measurable socioeconomic differences between the development and validation cohorts, and both cohorts are from a single country^10^.

In conclusion, a universal cervical-ultrasound screen for spontaneous preterm birth was not achievable across cohort and machine, but a high-risk subgroup showed a promising, if preliminary, signal. We believe the productive path forward is to predict distinct preterm-birth subtypes separately and to integrate additional biomarkers and data domains with imaging, rather than to pursue a single universal image-based screening model.

## Methods

### Study design and participants

We conducted a prediction-model development and external-validation study nested within two prospective observational pregnancy cohorts in north India, and report it following the TRIPOD+AI statement^23^. Models were developed and internally tested on the GARBH-Ini cohort at Gurugram Civil Hospital, Gurugram, Haryana^24^, and our best models were externally validated on the CORE cohort at the Hamdard Institute of Medical Sciences and Research (HIMSR), New Delhi; de-identified images were managed at the Translational Health Science and Technology Institute (THSTI), Faridabad.

Pregnant women were enrolled before 20 weeks of gestation with a dated ultrasound scan in GARBH-Ini, and at age over 18 years and before 20 weeks of gestation in CORE, with follow-up to delivery; the CORE cohort enrolled between August 2021 and March 2023. We included participants with an available mid-trimester (18 to 20 week) cervical ultrasound image and a recorded delivery outcome, and excluded multiple births, stillbirths, abortions, participants lost to follow-up with missing outcomes or scans, and, in GARBH-Ini, images that could not be retrieved through the de-identification pipeline.

We defined a clinically high-risk subgroup as participants with a history of previous pregnancy or preterm birth together with a short cervical length (<2.5 cm). All analyses were reported for the overall group and for this high-risk subgroup.

### Procedures and outcome

Cervical images were acquired following International Society of Ultrasound in Obstetrics and Gynecology guidance, on GE Voluson E8 and E8 Expert machines in GARBH-Ini and on a Samsung (HS70A) machine in CORE. Images were de-identified with an in-house tool; caliper annotations were removed by a cropping step and region-of-interest annotations by image inpainting, and the external set used clean (pre-caliper) images. Images were normalised using a data-specific mean and standard deviation and resized to 512 × 512 pixels for the texture pipeline. Data were partitioned at the participant level so that all images from a given participant were kept within a single split, with no leakage between development and test sets.

The predictors were the mid-trimester cervical image, a cervical-texture feature derived from it, cervical length measured at the 18 to 20 week scan, and 13 clinical variables (8 numeric and 5 categorical). The cervical region of interest, comprising the entire cervix, was manually annotated in the Computer Vision Annotation Tool by radiologists, and Local Binary Pattern texture (circular operator, radius 1, 16 sampling points, with bilinear interpolation) was computed within this region and summarised as a per-image histogram^8^. Missing clinical values were imputed using the median for numeric variables and the mode for categorical variables; several clinical variables were not collected in the CORE cohort and were treated as missing for the externally validated models.

The outcome was spontaneous preterm birth, a live birth before 37+0 weeks of gestation, with a secondary threshold of 34 weeks. Gestational age was established from the crown to rump length measured at enrolment using the Hadlock formula, and the outcome was taken from the recorded delivery date, independent of the imaging predictors.

### Model development

We developed three complementary families of models: classical machine-learning models using structured clinical variables, deep-learning models using the cervical ultrasound images, and multimodal models integrating both.

For the clinical-only models, numeric predictors were scaled with a robust scaler and categorical predictors were one-hot encoded. We trained logistic regression, random forest, support-vector machine, and AdaBoost classifiers. Hyperparameters were tuned by grid search under stratified 10-fold cross-validation on the training set.

For the image-based models, we used a Vision Transformer architecture for binary classification. Training used the AdamW optimiser with decoupled weight decay and a OneCycle learning-rate schedule. Class imbalance was addressed with data augmentation, focal loss (α = 0.9, γ = 2), and weighted random sampling. Because multiple frames were available per participant, image-level probabilities were aggregated to a participant-level score using the maximum predicted probability, applied consistently across test sets.

As a handcrafted alternative, rotation-invariant uniform Local Binary Pattern features (radius 1, P = 16) were extracted within the manually annotated cervical region and summarised as 59-bin histograms^8^. A Random Forest fitted on these texture histograms served as the primary texture model.

For multimodal prediction, Vision Transformer image features and the processed clinical features were combined by late fusion. The two representations were concatenated and passed through fully connected layers to produce the final probability, preserving modality-specific representations before integration. Class imbalance in the tabular and multimodal pipelines was additionally addressed with synthetic minority over-sampling (SMOTE) where needed.

### Statistical analysis

All models were evaluated on identical internal and external test sets. The primary measure of model performance was the area under the receiver-operating-characteristic curve (AUROC), reported together with sensitivity, specificity, positive predictive value, and negative predictive value. Classification thresholds other than 0.5 were selected using threshold-performance plots. Performance was reported separately for the overall group and the predefined high-risk subgroup, and at both the 37-week and 34-week thresholds.

Confidence intervals (95%) were obtained from 1,000 non-parametric bootstrap replicates, with stratified resampling that preserved outcome prevalence for the low-event analyses. No correction for multiple comparisons was applied across the grid of models, subgroups, and thresholds, which we therefore interpret as model selection and hypothesis generation rather than confirmatory testing. The study size was pragmatic, comprising all eligible participants with quality-passing images; no formal sample-size calculation was performed. Because preterm events were few, particularly within the high-risk subgroup and at the 34-week threshold, the number of events per candidate predictor was low and the internal estimates are likely to be optimistic.

## Data Availability

All data produced in the present study are available upon reasonable request to the authors

## Ethics approval and consent to participate

The study obtained the necessary ethics approvals from the institutional ethics committees of the Translational Health Science and Technology Institute, Faridabad (THS 1.8.1/(89)), Gurugram Civil Hospital, Gurugram (ECR/278/INST/HR/2013), and the Hamdard Institute of Medical Sciences and Research, New Delhi (HIMSR/IEC/43/2021). All methods were performed in accordance with the relevant guidelines and regulations and with the tenets of the Declaration of Helsinki, as part of the overall GARBH-Ini cohort objectives. Written informed consent was obtained from all participants after the nature of the study was explained. If an eligible woman was illiterate, a thumb impression was taken from her after ensuring that she had understood and had also explicitly stated her consent verbally, and a literate impartial witness signed the consent form.

## Acknowledgements

Professor M. K. Bhan will always be remembered reverently for his critical scientific and technical feedback. We greatly appreciate the extraordinary efforts from Drs. A Gambhir and S Sinha from DBT-India for supporting the GARBH-Ini program. We recognise the efforts of the research physicians, study nurses, clinical and laboratory technicians, field workers, internal quality improvement team, and the project and data management team of the GARBH-Ini cohort. We gratefully acknowledge our external radiology expert reviewers, Dr. Ritu Misra, Dr. Sandeep Bajaj, Dr. Gaurika Sahi, and Dr. Chanchal Singh, who reviewed ultrasound images monthly and helped maintain quality assurance throughout the study. The study figures were prepared by the authors. This work was funded by the DBT Wellcome Trust India Alliance (IA/CPHE/18/1/503947) and the Department of Biotechnology, Ministry of Science and Technology, Government of India (BT/PR9983/MED/97/194/2013).

The funders had no role in study design, data collection, analysis, interpretation, or the writing of this report. Members of GARBH-Ini (in alphabetical order): Translational Health Science and Technology Institute, NCR Biotech Cluster, Faridabad, India (Coordinating Institute): Shinjini Bhatnagar (PI), Vineeta Bal, Bhabatosh Das, Mahadev Dash, Bapu Koundinya Desiraju, Pallavi Kshetrapal, Sumit Misra, Uma Chandra Mouli Natchu, Satyajit Rath, Kanika Sachdeva, Dharmendra Sharma, Amanpreet Singh, Shailaja Sopory, Ramachandran Thiruvengadam, Nitya Wadhwa; National Institute of Biomedical Genomics, Kalyani, West Bengal, India: Arindam Maitra, Partha P Majumder (Co-PI), Souvik Mukherjee; Regional Centre for Biotechnology, NCR Biotech Cluster, Faridabad, India: Tushar K Maiti; Clinical Development Services Agency, Translational Health Science and Technology Institute, NCR Biotech Cluster, Faridabad, India: Monika Bahl, Shubra Bansal; Gurugram Civil Hospital, Haryana, India: Umesh Mehta, Sunita Sharma, Brahmdeep Sindhu; Safdarjung Hospital, New Delhi, India: Sugandha Arya, Rekha Bharti, Harish Chellani, Pratima Mittal; Maulana Azad Medical College, New Delhi, India: Anju Garg, Siddharth Ramji; The Ultrasound Lab, Defence Colony, New Delhi, India: Ashok Khurana; Hamdard Institute of Medical Sciences and Research, Jamia Hamdard University, New Delhi, India: Reva Tripathi; All India Institute of Medical Sciences, New Delhi, India: Alpesh Goyal, Yashdeep Gupta, Smriti Hari, Nikhil Tandon; Government of Haryana, India: Rakesh Gupta; International Centre for Genetic Engineering and Biotechnology, New Delhi, India: Dinakar M Salunke (Co-PI); G Balakrish Nair (Rajiv Gandhi Centre for Biotechnology, Trivandrum); Gagandeep Kang (Christian Medical College, Vellore).

## Author contributions

B.K.D. and S.B. conceived and supervised the study. D.M. and R.C. analysed the data and built the artificial-intelligence models. N.S., R.J., and R.C. performed the statistical analysis. A.K. advised on imaging and ultrasound protocols and provided critical review of the manuscript. R.Tr. and A.J. provided clinical and radiological expertise and oversaw imaging and outcome data collection at the HIMSR validation site. S.B., J.A.N., R.Th., N.W., and B.K.D. guided and provided feedback on the analysis and interpretation of the results. R.C., D.M., R.J., B.K.D., R.Th., and S.B. wrote the paper. All authors reviewed and approved the manuscript.

Initials: R.C., Radhika Chanian; D.M., Divyanshu Mishra; R.J., Rahul Jain; N.S., Nikhil Sharma; A.K., Ashok Khurana; R.Tr., Reva Tripathi; A.J., Abhinav Jain; N.W., Nitya Wadhwa; J.A.N., J. Alison Noble; R.Th., Ramachandran Thiruvengadam; S.B., Shinjini Bhatnagar; B.K.D., Bapu Koundinya Desiraju.

## Data availability

The data described in the manuscript will be made available upon request pending approval. The analysis code will be made available upon request pending approval. Cervical ultrasound images are sensitive de-identified data stored on a THSTI server; the terms of any sharing will be specified accordingly.

