## Supplementary Material for "Development and external validation of deep learning models for spontaneous preterm birth prediction from mid-trimester cervical ultrasound"

### **S1. Study overview and elements common to all models**

#### **S1.1 Prediction task, cohorts, and outcome**

The outcome was spontaneous preterm birth (sPTB), defined as a live birth before 37+0 weeks of gestation (primary threshold), with a secondary threshold of before 34+0 weeks. Gestational age was established from the crown–rump length at enrolment using the Hadlock formula, and the outcome was taken from the recorded delivery date, independent of the imaging predictors. Every model was developed and internally tested on the prospective GARBH-Ini cohort (US images obtained from GE Voluson E8 and E8 Expert machines) and, for the best-performing configurations, externally validated on the independent CORE/Jamia cohort (scanned on a Samsung HS70A machine). The change of both cohort and ultrasound vendor between development and validation makes the external test a demanding assessment of generalisation.

#### **S1.2 Groups and thresholds**

All analyses were reported for two groups and, where events permitted, at two gestational-age thresholds:

- **Overall group —** all eligible participants with a quality-passing mid-trimester (18–20 week) cervical scan and a recorded delivery outcome.
- **Clinically high-risk subgroup —** participants with a history of previous pregnancy or previous preterm birth together with a short cervical length (< 2.5 cm). This subgroup is small, particularly at the 34-week threshold, so its estimates are imprecise and treated as exploratory.
- **Thresholds —** the < 37-week definition is primary; the < 34-week definition is secondary and rests on fewer preterm events.

#### **S1.3 Image de-identification and partitioning**

Cervical images were acquired following ISUOG guidance and de-identified with an in-house tool: caliper annotations were removed by a cropping step and region-of-interest annotations by image inpainting; the external set used clean (pre-caliper) images. Data were partitioned strictly at the participant level, so that all images from a given participant remained within a single split, preventing leakage between development and test sets. Because multiple images were available per participant for the image-based models, image-level probabilities were aggregated to a single participant-level score using the maximum predicted probability, applied consistently across the internal and external test sets.

#### **S1.4 Clinical predictors and missing-data handling**

The structured predictors comprised cervical length measured at the 18–20-week ultrasound examination and 13 clinical variables (8 numeric and 5 categorical). The categorical variables were safe cooking fuel, consanguinity, history of recurrent abortion, number of previous preterm births, and a composite high-risk pregnancy indicator. Missing values were imputed using the median for numeric variables and the mode for categorical variables. Imputation statistics were estimated using the training set only and applied without modification to the validation, internal test, and external test sets to prevent data leakage. Several clinical variables were unavailable in the CORE cohort and were therefore treated as missing during external validation.

### **S2. Machine-learning models on structured clinical variables**

The first family used only structured, tabular information (clinical variables, optionally with cervical length) and classical supervised classifiers. These models establish how much predictive signal is available from routinely recorded clinical data before any imaging is added.

#### **S2.1 Feature preprocessing**

- **Imputation —** median for numeric predictors, mode for categorical predictors, fit on the training split only.
- **Scaling —** numeric predictors were transformed with a RobustScaler, since it limits the influence of outliers common in clinical measurements.
- **Encoding —** categorical predictors were one-hot encoded; multi-level history variables (e.g., number of previous preterm children) expanded into their corresponding indicator columns.
- **Class balancing —** the strong term/preterm imbalance in the training data was addressed with the Synthetic Minority Over-sampling Technique (SMOTE), applied to the training partition only, after the train/validation split, so that no synthetic samples leaked into evaluation.

#### **S2.2 Classifiers and hyperparameter tuning**

Four classifiers were trained and compared: logistic regression, random forest, support-vector machine, and AdaBoost. Hyperparameters were selected by exhaustive grid search under stratified 10-fold cross-validation on the training set, optimising the cross-validated AUROC. For AdaBoost (the SAMME variant), the grid spanned the number of estimators (50–80) and the learning rate (0.001–0.1); for the tree- and margin-based learners the grids covered the corresponding depth, estimator-count, and regularisation parameters. The best cross-validated estimator was refitted and carried forward to the held-out internal test set and, where applicable, the external test set.

#### **S2.3 Experiments and key results**

Two structured-data configurations were evaluated across both groups and both thresholds: clinical variables alone, and clinical variables augmented with cervical length. In the overall group these models were modest at the 37-week threshold (internal AUROC ≈ 0.61) and did not generalise externally (≈ 0.51–0.52). The clearest difference emerged in the high-risk subgroup at the 34-week threshold, where the clinical-variable model reached an internal AUROC of 0.77 (95% CI 0.53–0.95) and an external AUROC of 0.86 (95% CI 0.75–0.98), and the clinical-plus-cervical-length model reached 0.80 (0.55–0.95) internally and 0.85 (0.69–0.96) externally (Refer to Table 3). These high-risk estimates rest on very few preterm events (as few as three externally), so their confidence intervals are wide, and several operating points are degenerate; they are reported for transparency and interpreted as hypothesis-generating. In detail point estimates and intervals appear in Supplementary Table S1.

### **S3. Deep-learning models on cervical ultrasound images**

The second family learned directly from the maternal cervical ultrasound images using a Vision Transformer (ViT) architecture. The aim was to test whether an end-to-end image model could extract preterm-related signal beyond that available from cervical length and clinical variables.

#### **S3.1 Architecture and inputs**

- **Backbone —** a ViT-Base with 32×32 patches at 384-pixel resolution (vit_base_patch32_384), initialised from ImageNet-pretrained weights and adapted to a single input channel (greyscale ultrasound) with a single-logit binary-classification head.
- **Input pipeline —** images were converted to greyscale, resized to 384×384, and normalised with a dataset-specific mean (0.2782) and standard deviation (0.2063).
- **Participant-level scoring —** per-image probabilities were aggregated to one score per participant using the maximum predicted probability.

#### **S3.2 Optimisation**

Training used the AdamW optimiser (initial learning rate 1×10⁻³, weight decay 1×10⁻²) with decoupled weight decay, under a OneCycle learning-rate schedule (peak learning rate 1×10⁻²). Models were trained with mixed (16-bit) precision and a batch size of 64, with checkpoints selected on the highest validation AUROC. To stabilise estimates, each configuration was trained across five independent runs.

#### **S3.3 Class-imbalance strategy and ablation**

Because preterm cases are rare, class imbalance was central to training. The core loss was a sigmoid focal loss (α = 0.9, γ = 2), which down-weights easy, well-classified majority examples and focuses learning on the hard minority class. On top of this loss, two complementary imbalance mechanisms were ablated: weighted random sampling, which over-samples the minority class within each mini-batch, and data augmentation via random horizontal flipping. The four resulting configurations are summarised below.

| **Configuration** | **Focal loss** | **WRS** | **Augmentation** | **Role in the ablation** |
| --- | --- | --- | --- | --- |
| **Focal Loss** | α=0.9, γ=2 | — | — | Baseline: loss-based re-weighting only (shuffled sampling). |
| **Focal Loss + Data Augmentation** | α=0.9, γ=2 | — | Horizontal-flip | Adds input variability to reduce overfitting. |
| **Focal Loss + Weighted Random Sampler** | α=0.9, γ=2 | ✓ | — | Adds minority over-sampling at the batch level. |
| **Focal Loss + Weighted Random Sampler + Data Augmentation** | α=0.9, γ=2 | ✓ | Horizontal-flip | Combined strategy carried forward as the reported ViT model. |

**Supplementary Table S2.** *Vision Transformer class-imbalance ablation. All four variants share the ViT-Base/32-384 backbone, single-channel input, focal loss, AdamW + OneCycle optimisation, and five-run training; they differ only in the sampling and augmentation strategy layered on top of the focal loss.*

#### **S3.4 Key results**

In the overall group the ViT was close to chance internally at the 37-week threshold (AUROC 0.51, 95% CI 0.46–0.57) and modest externally (0.56, 0.49–0.65); at the 34-week threshold it reached 0.54 (0.46–0.60) internally and 0.59 (0.49–0.69) externally. In the high-risk subgroup at 37 weeks, it reached 0.56 (0.47–0.65) internally and 0.57 (0.45–0.68) externally. The Vision Transformer did not outperform the cervical-texture model on either test set. All point estimates are given in Supplementary Table S1.

### **S4. Multimodal model**

The third family combined the imaging and clinical data streams to test whether jointly modelling both improved predictions over either modality alone. The design used late fusion methodology: each modality was encoded separately, and the two representations were concatenated only just before the fully connected layer, preserving modality-specific structure before integration.

#### **S4.1 Architecture**

- **Image branch —** the same ViT-Base/32-384 backbone (single-channel input) as in S3, used here as a feature extractor with its classification head removed; the class-token embedding (768-dimensional) served as the image representation.
- **Clinical branch —** the processed clinical vector was passed through a projection block: a linear layer to 128 dimensions, ReLU activation, batch normalisation, and dropout (p = 0.2).
- **Fusion and head —** the 768-dimensional image embedding and the 128-dimensional clinical embedding were concatenated (896 dimensions) and passed through a classifier: linear layer to 256 dimensions, ReLU, dropout (p = 0.3), and a final single-logit layer.

#### **S4.2 Clinical preprocessing for fusion**

For the multimodal pipeline the clinical features were processed with a scikit-learn column transformer: numeric predictors (e.g., interval since last delivery and derived BMI) were median-imputed and standardised (zero mean, unit variance), while categorical predictors (safe fuel, consanguinity, recurrent abortion, previous preterm children, high-risk-pregnancy flag) were mode-imputed and one-hot encoded. The transformer was fit on the training partition only and applied unchanged to validation and test data. Residual class imbalance in the tabular and multimodal pipelines was additionally addressed with SMOTE where needed.

#### **S4.3 Optimisation**

The multimodal model was trained with the same sigmoid focal loss (α = 0.9, γ = 2) and AdamW optimiser (learning rate 1×10⁻³, weight decay 1×10⁻²) under a OneCycle schedule (peak 1×10⁻²), with mixed-precision training, a batch size of 84, and checkpoint selection on validation AUROC across five independent runs. Participant-level aggregation followed the same maximum-probability rule.

#### **S4.4 Key results**

Fusion did not provide us a benefit over the single-modality models. In the overall group at 37 weeks the multimodal model reached an internal AUROC of 0.50 (95% CI 0.43–0.56) and an external AUROC of 0.47 (0.38–0.55); in the high-risk subgroup at 37 weeks, it reached 0.56 (0.44–0.67) internally and 0.54 (0.41–0.69) externally. These values are reported alongside all other confidence intervals in Supplementary Table S1.

### **S5. Cervical-texture model (Local Binary Patterns + Random Forest)**

The fourth family is a handcrafted, interpretable texture pipeline motivated by prior work associating quantitative cervical-ultrasound texture with sPTB. It was the best-performing image-based model on internal testing and is the model featured in the main-text ROC figure and operating-point table.

#### **S5.1 Region-of-interest annotation**

The cervical region of interest, comprising the entire cervix, was manually annotated by radiologists in the Computer Vision Annotation Tool (CVAT). Texture features were then computed only within this masked region, so that the descriptor reflects cervical tissue rather than surrounding structures. For processing, each image was read, resized to 512×512 pixels, and converted to greyscale; the binarised mask was used both to restrict the texture computation and to build the region overlay.

#### **S5.2 Local Binary Pattern descriptor**

- **Operator —** a circular Local Binary Pattern with radius 1 and P = 16 sampling points; neighbour intensities were obtained by bilinear interpolation at the sampled angular positions.
- **Uniform mapping —** each 16-bit pattern was tested for uniformity (at most two 0↔1 transitions around the circle). Uniform patterns retained their identity while all non-uniform patterns were collapsed into a single additional bin, yielding a compact, rotation-tolerant, noise-robust descriptor.
- **Feature vector —** the LBP responses inside the cervical mask were summarised as a 59-bin histogram, giving one fixed-length texture vector per image.

#### **S5.3 Classifier, balancing, and tuning**

A Random Forest model was fitted on the texture histograms. Class imbalance was addressed with SMOTE on the training partition, and hyperparameters were tuned by grid search under stratified 10-fold cross-validation optimising AUROC, spanning the number of trees (50–90) and maximum depth (3–6). Three model configurations were evaluated: (1) cervical texture features within the ROI alone, (2) cervical texture features combined with cervical length, and (3) cervical texture features combined with cervical length and clinical variables. This comparison assessed whether cervical texture provides predictive information beyond that captured by cervical length and clinical variables.

#### **S5.4 Key results**

The best-performing image model on internal testing was the cervical-texture Random Forest. In the overall group it reached an internal-test AUROC of 0.71 (95% CI 0.60–0.82; 94 images: 12 preterm, 82 term) but fell to 0.52 (95% CI 0.38–0.64; 69 images: 33 preterm, 36 term) (Refer to Table 2) on the external cohort discrimination close to chance after the change of cohort and machine. Adding cervical length or clinical variables to the texture features did not improve overall discrimination (LBP ROI + cervical length: internal 0.61, external 0.50; LBP ROI + cervical length + clinical: internal 0.61, external 0.51).

### **S6. Evaluation and statistical analysis**

All models were assessed on identical internal and external test sets. The primary performance measure was the area under the receiver-operating-characteristic curve (AUROC), reported together with sensitivity, specificity, positive predictive value, and negative predictive value. Classification thresholds were selected from threshold-performance curves to achieve an appropriate balance between sensitivity and specificity, and metrics were computed at the selected threshold. Performance was reported separately for the overall cohort and the predefined high-risk subgroup, and for both the <37-week and <34-week preterm birth endpoints.

95% confidence intervals were obtained from 1,000 non-parametric bootstrap replicates, with stratified resampling that preserved outcome prevalence in the low-event analyses. No correction for multiple comparisons was applied across the grid of models, subgroups, and thresholds; the exercise is therefore interpreted as model selection and hypothesis generation rather than confirmatory testing. Because preterm events were few, particularly within the high-risk subgroup and at the 34-week threshold, the number of events per candidate predictor was low, and the internal estimates are likely to be optimistic. Given the low positive predictive values observed at these prevalences, none of the models is suitable for standalone clinical use in its current form.

### **Supplementary Table S1. AUROC by model, group, and threshold**

| **Experiment** | **Group** | **PTB def.** | **Internal AUROC (95% CI)** | **External AUROC (95% CI)** |
| --- | --- | --- | --- | --- |
| **Clinical variables** | Overall | <37 | 0.61 (0.55, 0.67) | 0.52 (0.46, 0.60) |
| **Clinical variables** | Overall | <34 | 0.65 (0.51, 0.80) | 0.68 (0.44, 0.86) |
| **Clinical variables** | High risk | <37 | 0.50 (0.39, 0.60) | 0.62 (0.38, 0.81) |
| **Clinical variables** | High risk | <34 | 0.77 (0.53, 0.95) | 0.86 (0.75, 0.98) |
| **Clinical + cervical length** | Overall | <37 | 0.61 (0.55, 0.67) | 0.51 (0.44, 0.60) |
| **Clinical + cervical length** | Overall | <34 | 0.76 (0.64, 0.87) | 0.59 (0.37, 0.80) |
| **Clinical + cervical length** | High risk | <37 | 0.54 (0.46, 0.63) | 0.46 (0.24, 0.68) |
| **Cervix US images (ViT)** | Overall | <37 | 0.51 (0.46, 0.57) | 0.56 (0.49, 0.65) |
| **Cervix US images (ViT)** | Overall | <34 | 0.54 (0.46, 0.60) | 0.59 (0.49, 0.69) |
| **Cervix US images (ViT)** | High risk | <37 | 0.56 (0.47, 0.65) | 0.57 (0.45, 0.68) |
| **LBP ROI + cervical length** | Overall | <37 | 0.61 (0.55, 0.68) | 0.50 (0.44, 0.56) |
| **LBP ROI + CL + clinical** | Overall | <37 | 0.61 (0.55, 0.66) | 0.51 (0.45, 0.59) |
| **Multimodal** | Overall | <37 | 0.50 (0.43, 0.56) | 0.47 (0.38, 0.55) |
| **Multimodal** | High risk | <37 | 0.56 (0.44, 0.67) | 0.54 (0.41, 0.69) |

**Supplementary Table S1.** *AUROC discrimination across the full grid of clinical, deep-learning, texture-augmented, and multimodal configurations. Values are reproduced from the source results table; CL = cervical length; LBP = Local Binary Patterns; ROI = region of interest; ViT = Vision Transformer.*
